# A Methodological Framework for Passive Visual Mirror Exposure in Children with CVI and an Atelic Profile

**DOI:** 10.64898/2026.01.08.26343692

**Authors:** Edwige Smague

## Abstract

Children with cerebral visual impairment (CVI) presenting an atelic profile constitute a population that is largely excluded from neurovisual research, primarily due to the absence of reliable behavioural responses to standard assessment or intervention paradigms. This exclusion often leads to the assumption that a lack of observable response reflects an absence of visual processing, despite converging evidence from neuroscience indicating that neural processing may persist in the absence of overt behavioural expression.

This article proposes a pre-empirical methodological framework describing **a** passive visual mirror exposure paradigm intended to inform the design of future feasibility studies in children with CVI and an atelic profile. The visual mirror is defined as the repeated presentation of a highly predictable, coherent visual experience, intended to reduce perceptual instability and cognitive load associated with typical visual environments, without aiming to restore or train visual function.

The paradigm relies on passive visual exposure delivered via a large screen, or, where appropriate and tolerated, a virtual reality headset configured in a strictly non-interactive mode, without requiring effort, task engagement, or voluntary responses from the child. The proposed framework is grounded in a precautionary ethical rationale specifically adapted to behaviourally non-responsive paediatric populations, defined here as children presenting absent voluntary behavioural responses to sensory or task-based instructions.

The framework outlines predefined safety criteria and non-verbal physiological indicators, including heart rate and autonomic markers, as illustrative examples of feasibility and tolerability monitoring, rather than behavioural performance.

This manuscript does not report or initiate research involving human participants. It is intended to characterise the ethical acceptability, conceptual implementability, and methodological considerations of a passive visual mirror paradigm in this vulnerable population, and to inform future research considerations contingent upon formal ethical approval, without implying any obligation or commitment to subsequent studies.

## 1. Introduction

Cerebral visual impairment (CVI) (1) is now recognised as one of the leading causes of paediatric visual disability. It results from damage to or dysfunction of the cerebral pathways involved in visual processing, independent of ocular integrity (2). Visual difficulties associated with CVI are heterogeneous and may include impairments in gaze orienting, motion perception, object recognition, sustained visual attention, or visuomotor coordination.

Within this population, children presenting an atelic profile constitute a particularly marginalised group. Atelicity is defined here as a persistent inability to initiate, maintain, or orient behaviour towards a goal, even in simple sensory contexts (3). Children with atelic profiles are typically unable to follow instructions or produce reliable behavioural responses, which effectively excludes them from conventional visual assessment and rehabilitation paradigms that rely on task engagement or observable performance.

This exclusion has created a significant methodological and ethical gap. The absence of empirical data in this subgroup makes it difficult to determine whether these children are genuinely non-responsive to visual stimuli or whether they are simply inaccessible to existing assessment methods. In this context, behavioural non-responsiveness cannot be assumed to reflect an absence of visual processing, as converging evidence from neuroscience indicates that perceptual and neural processing may persist in the absence of overt behavioural expression.

The present paper addresses this gap by proposing a feasibility-informed methodological framework designed to conceptually examine physiological responses to controlled visual stimuli in the absence of voluntary behaviour. Rather than aiming to assess therapeutic efficacy, clinical benefit, or functional visual improvement, the framework is explicitly limited to outlining methodological and ethical conditions under which a passive visual mirror exposure paradigm could be evaluated in children with CVI and an atelic profile, while allowing the acquisition of stable, non-behavioural physiological signals.

Accordingly, the primary objective of this work is not to report an implemented study, but to define and formalise the methodological and ethical considerations relevant to determining whether repeated sessions of passive visual mirror exposure may be feasible and tolerable under controlled conditions in behaviourally non-responsive paediatric populations. The central feasibility question is therefore presented as a prospective and conceptual inquiry, rather than as an empirically tested outcome.

## 2. The Atelic Child and Visual Experience

### 2.1 Atelicity as a Disruption of the Perception–Prediction-Action Loop

In children presenting an atelic profile, the absence of goal-directed behaviour should not be interpreted as an absence of perception. Evidence from cognitive and neuroscientific research indicates that perceptual and memory-related processing can occur in the absence of voluntary control or overt behavioural expression (4). Accordingly, limited behavioural responsiveness does not preclude the presence of underlying sensory processing.

Atelicity can be described as a functional difficulty in initiating, maintaining, or organising behaviour in response to sensory information. In this context, the child may receive sensory input but be unable to use it reliably to guide action, exploration, or sustained engagement with the environment (5). This description does not imply a deficit in sensory reception itself, but rather a disruption in the translation of sensory information into observable, goal-directed behaviour.

### 2.2 Visual Experience in Atelic Children with Cerebral Visual Impairment

In atelic children with cerebral visual impairment, visual experience is often reported by clinicians and caregivers as unstable and difficult to interpret (6). Commonly described features include:

- **Image Instability**: where the visual scene may appear fragmented or inconsistent;
- **Excessive Perceptual Detail**: resulting in difficulty prioritising or organising visual information;
- **Difficulty Anticipating Visual Change**: such that new objects or changes in lighting may be experienced as abrupt or unpredictable;
- **Impaired Recognition of Visual Stability**: for example, difficulty recognising that an object remains the same despite minor changes in position or context.

At a functional level, these characteristics are associated with increased perceptual uncertainty and elevated cognitive load during visual processing. Over time, visual engagement may become effortful or aversive, leading to reduced spontaneous visual exploration or apparent disengagement from visual stimuli (7).

This section provides a conceptual and descriptive framework to support the design choices of the proposed protocol. It does not constitute a testable theoretical model and does not imply specific mechanisms of impairment. Rather, it serves to justify the use of simplified, predictable, and non-interactive visual stimuli within a pre-empirical methodological framework involving behaviourally non-responsive paediatric populations.

## 3. Research Question and Objectives

### 3.1 Primary Feasibility Question

The visual mirror exposure paradigm described in this article is conceptually informed by principles derived from mirror-based approaches originally developed in post-stroke motor rehabilitation (8), while differing fundamentally in its intended function and mechanism. Rather than relying on visual comparison or illusion, the present paradigm is designed to provide a coherent, stable, and highly predictable visual stimulus that may temporarily substitute for unstable or cognitively costly visual environments in children with cerebral visual impairment, consistent with sensory substitution principles (9) and descriptive models of predictive processing (7).

In the context of cerebral visual impairment and atelic profiles, the paradigm is explicitly designed for behaviourally non-responsive paediatric populations and does not require task engagement, instruction following, or voluntary behavioural responses.

The primary feasibility question articulated in this manuscript is therefore formulated as a prospective and conceptual inquiry, as follows: Would it be feasible to deliver repeated sessions of passive visual mirror exposure to children with cerebral visual impairment and an atelic profile while maintaining physiological tolerability and obtaining stable, interpretable non-behavioural physiological signals in behaviourally non-responsive paediatric populations?

### 3.2 Primary Objective

The primary objective of this work is to define and formalise the feasibility and physiological tolerability criteria relevant to a passive visual mirror exposure protocol in the context of future empirical studies involving children with cerebral visual impairment and an atelic profile. Feasibility is defined in terms of the theoretical implementability of the protocol without inducing sustained physiological distress, and while enabling the potential acquisition of usable autonomic physiological measures in a population for whom active, task-based paradigms are not accessible (7).

### 3.3 Secondary Methodological Objectives

The secondary objectives outlined here are strictly methodological and illustrative. They are intended to characterise the types of protocol performance indicators and measurement properties that may be relevant rather than to assess therapeutic efficacy or clinical benefit. Specifically, the objectives are:

- to assess whether non-behavioural physiological measures, including heart rate and heart rate variability, could in principle be reliably recorded during passive visual mirror exposure sessions in this population;
- to examine the intra-individual stability and variability of these physiological signals as a methodological consideration under conditions of highly controlled and predictable visual stimulation (7);
- to outline potential rates of session completion, interruption, or early termination due to indicators of non-verbal dissent or physiological distress, in accordance with precautionary ethical principles;
- to explore whether protocol tolerability may reasonably be expected to vary across individual neurovisual profiles within the atelic population, without making claims of group-level inference.

### 3.4 Reference to Feasibility, Tolerability and Safety Criteria

To ensure the highest standard of participant safety, the present manuscript specifies feasibility and tolerability criteria as a pre-empirical governance framework. These predefined thresholds and decision rules, detailed in Table 1, constitute a conceptual safety and oversight structure intended to guide future protocol implementation.

**Table 1.**
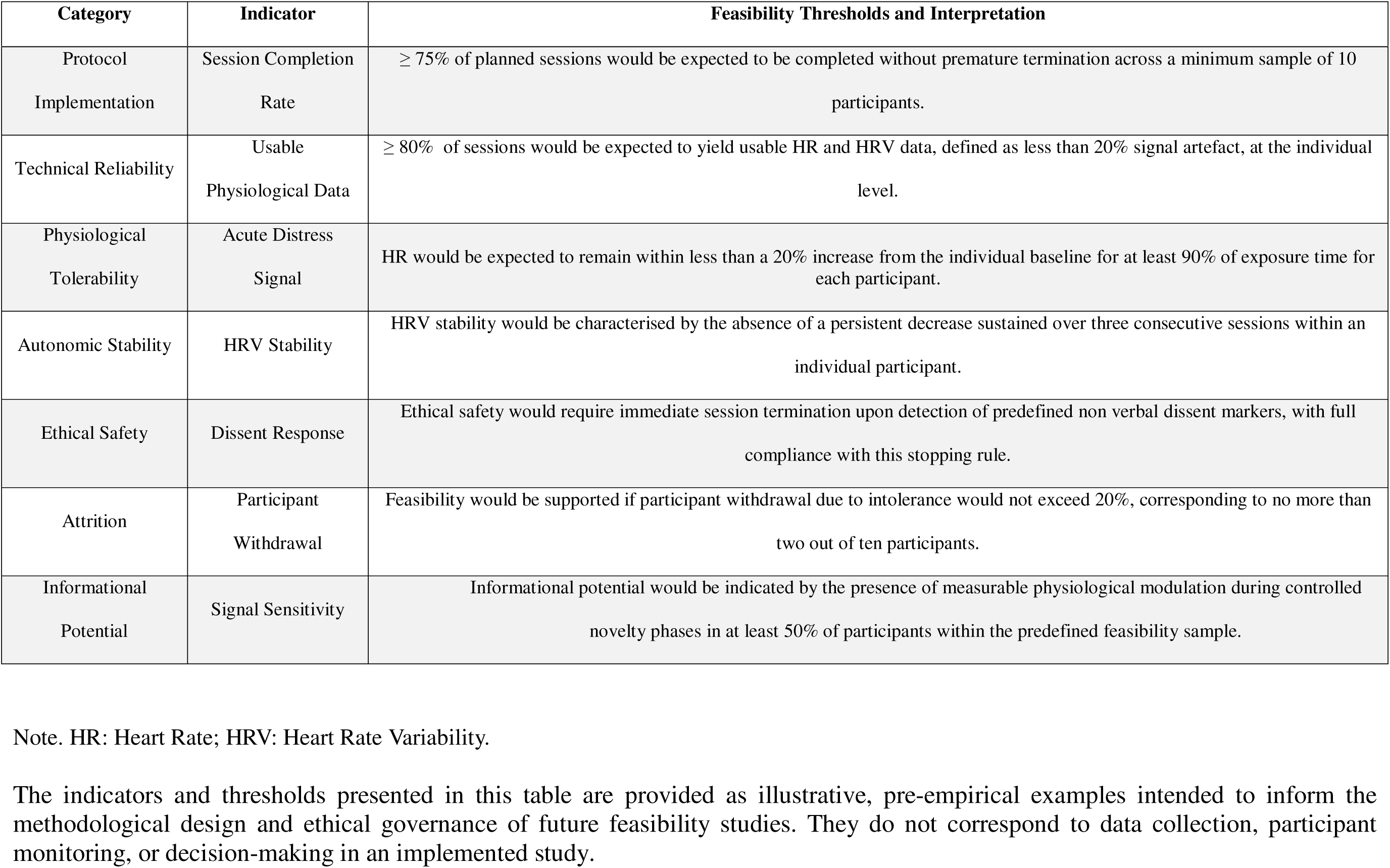
Illustrative feasibility, tolerability, and safety indicators with predefined decision thresholds (pre-empirical framework).

| Category | Indicator | Feasibility Thresholds and Interpretation |
| --- | --- | --- |
| Protocol Implementation | Session Completion Rate | ≥ 75% of planned sessions would be expected to be completed without premature termination across a minimum sample of 10 participants. |
| Technical Reliability | Usable Physiological Data | ≥ 80% of sessions would be expected to yield usable HR and HRV data, defined as less than 20% signal artefact, at the individual level. |
| Physiological Tolerability | Acute Distress Signal | HR would be expected to remain within less than a 20% increase from the individual baseline for at least 90% of exposure time for each participant. |
| Autonomic Stability | HRV Stability | HRV stability would be characterised by the absence of a persistent decrease sustained over three consecutive sessions within an individual participant. |
| Ethical Safety | Dissent Response | Ethical safety would require immediate session termination upon detection of predefined non verbal dissent markers, with full compliance with this stopping rule. |
| Attrition | Participant Withdrawal | Feasibility would be supported if participant withdrawal due to intolerance would not exceed 20%, corresponding to no more than two out of ten participants. |
| Informational Potential | Signal Sensitivity | Informational potential would be indicated by the presence of measurable physiological modulation during controlled novelty phases in at least 50% of participants within the predefined feasibility sample. |

Their primary purpose is to characterise the reliability, tolerability, and ethical acceptability of the protocol in children with atelic profiles, and to ensure that any future empirical study remains grounded in continuous monitoring of participant well-being and protocol stability. These criteria are not intended to demonstrate therapeutic efficacy but to guide ethical conduct and interpretation within a feasibility-focused study.

Table 1 summarises the primary feasibility question, objectives, indicators, and predefined feasibility and tolerability criteria as part of a pre-empirical methodological framework. All criteria are specified a priori to illustrate how ethical conduct, safety monitoring, and outcome interpretation could be structured in future feasibility studies, rather than to report procedures applied in an implemented study. The table links each research objective to concrete physiological or procedural indicators and to explicit decision rules intended to guide prospective protocol design and ethical governance, rather than protocol continuation or early termination in an ongoing study. Importantly, these criteria are not intended to demonstrate therapeutic efficacy or clinical benefit. Rather, they are designed to conceptually examine whether the proposed visual mirror exposure protocol may be considered ethically manageable and methodologically implementable under controlled conditions in children with cerebral visual impairment and an atelic profile, while enabling the potential acquisition of interpretable non-behavioural physiological data. This structure supports transparency, ethical oversight, and methodological clarity at the protocol design stage, without implying any obligation, initiation, or commitment to subsequent research.

## 4. Methods

### 4.1 Participants

The target population comprises children with cerebral visual impairment (CVI) presenting an atelic profile, defined as a persistent inability to initiate, maintain, or orient behaviour towards a goal, even in simple sensory contexts. This population is characterised as behaviourally non-responsive, which currently precludes participation in conventional visual assessment or rehabilitation protocols that rely on task engagement or observable performance.

No participants are recruited as part of the present manuscript. Any future empirical implementation of the proposed protocol would require prior approval from an appropriate Human Research Ethics Committee and would be conducted within an institutional framework.

Given the conceptual and pre-empirical scope of this work, no formal sample size calculation is performed. A minimum target sample of 10 participants is specified here illustratively, as a commonly accepted benchmark in feasibility study design, to indicate the scale at which protocol feasibility, tolerability, and measurement reliability could be evaluated in this rare and heterogeneous population using an intra-individual design.

#### Inclusion Criteria

In a prospective implementation scenario, participants would be expected to meet the following criteria:

- A clinical diagnosis of cerebral visual impairment, established by a qualified clinician and independent of ocular pathology;
- A behavioural profile consistent with atelicity, operationalised as the absence of reliable goal-directed responses to visual or task-based instructions;
- Age within the paediatric range appropriate for non-invasive physiological monitoring, as would be determined by the host institution’s ethics committee;
- Medical stability permitting participation in brief passive visual exposure sessions without contraindication;
- Informed consent would be obtained from a parent or legal guardian.

#### Exclusion Criteria

In a future empirical context, participants would be expected to be excluded if any of the following criteria were met:

- Uncontrolled epilepsy or recent seizure activity judged to pose an increased risk during visual stimulation;
- Severe medical instability or acute illness at the time of participation;
- Known intolerance to screen-based or head-mounted visual exposure;
- Conditions preventing reliable acquisition of physiological signals, such as severe movement disorders incompatible with sensor placement;
- Any clinical judgement indicating that participation would not be in the child’s best interest.

#### Ethical Justification

The ethical justification for the present protocol lies in its methodological design rather than solely in the general vulnerability of the target population. The proposed visual mirror exposure paradigm is strictly passive and non-invasive, and does not require instruction following, task engagement, or voluntary behavioural responses. As such, it is specifically adapted to children for whom conventional assessment or intervention procedures are not ethically appropriate due to their limited capacity to provide reliable behavioural assent.

Given the reduced capacity for verbal or behavioural communication in behaviourally non-responsive children with an atelic profile, the absence of overt response would not be interpreted as consent. Participant protection would be ensured through continuous monitoring of predefined physiological and behavioural indicators of potential non-verbal dissent. These indicators and their associated thresholds are specified a priori and operationalised in Table 1 as part of a pre-empirical ethical governance framework. In particular, session termination would be triggered by predefined autonomic criteria aligned with Table 1, including a sustained heart rate increase beyond the 20% threshold relative to the individual baseline, a persistent reduction in heart rate variability across consecutive sessions, and/or the emergence of behavioural markers indicative of distress or refusal.

Any detection of such indicators would result in immediate termination of the session, in accordance with the predefined stopping rules. This precautionary approach prioritises participant safety and minimises risk exposure while allowing conceptual evaluation of protocol feasibility under controlled conditions.

The present manuscript does not report or initiate a feasibility study. It does not involve therapeutic intent, does not seek to induce functional change, and does not withhold standard care. Its purpose is limited to defining the ethical and methodological conditions under which the protocol could be implemented safely and tolerably in this population, based on explicit, predefined safety thresholds, without implying any obligation or commitment to subsequent research.

### 4.2 Setting and Equipment

In a future implementation, the visual mirror exposure paradigm would be delivered in a controlled clinical or research setting designed to minimise extraneous sensory stimulation and to ensure participant safety. Sessions would take place in a quiet room with reduced ambient noise and simplified visual surroundings, in order to limit non-specific sensory load and potential sources of distraction. The protocol is designed to be implementable across clinical and research settings without requiring site-specific adaptation.

#### Modes of Visual Presentation

Visual stimuli would be presented using one of two non-interactive modalities, selected on the basis of participant tolerance, comfort, and clinical judgement:

- A large screen positioned directly in front of the child, which constitutes the default mode of visual presentation and would be used whenever head-mounted devices are not tolerated or are deemed inappropriate.
- A virtual reality headset, configured in a strictly passive, non-interactive mode, providing full-field visual exposure and reducing peripheral visual input. Use of a head-mounted device would be optional and contingent upon individual tolerance; it would be discontinued immediately if signs of discomfort or non-verbal dissent are observed.

Both modalities are designed to deliver identical visual content and temporal parameters. No interactive features, task demands, or response contingencies are included in either modality. The protocol is conceptually evaluated for feasibility across presentation modalities rather than for equivalence or comparative performance between modalities.

#### Controlled Conditions

Across both modes of visual presentation, the following parameters would be controlled and standardised:

- Stimulus size, contrast, luminance, colour, and motion characteristics.
- Duration of visual exposure blocks and inter-block intervals.
- Viewing distance in the screen condition, adjusted to promote central visual exposure while limiting peripheral engagement.
- Absence of additional visual, auditory, or tactile stimulation during exposure.

The purpose of this controlled environment is not to create an immersive or engaging experience, but to ensure precise regulation of visual input and to reduce non-specific sensory stimulation. This configuration is intended to approximate a mirror-like condition in which the child would be exposed to a single, isolated, and predictable visual stimulus under conditions of minimal sensory competition.

#### Equipment for Physiological Monitoring

Physiological data would be acquired using non-invasive sensors suitable for paediatric populations. Monitoring would focuses on autonomic measures, including heart rate and heart rate variability, that could be recorded continuously during baseline, exposure, and post-exposure periods.

All physiological monitoring devices would be required to be certified for human use and comply with applicable regulatory standards (e.g. CE marking and/or TGA or FDA approval, depending on device availability and site requirements). Detailed device specifications would be provided in supplementary documentation or appendices where required by the reviewing ethics committee.

Sensor placement and data acquisition procedures would be selected to minimise discomfort and to allow reliable signal recording without requiring voluntary movement, verbalisation, or task engagement from the child. Signal quality would be assessed in real time to identify artefacts resulting from excessive movement, sensor displacement, or technical failure. Periods of data affected by such artefacts would be flagged and excluded from analysis according to predefined criteria.

#### Safety Monitoring and Personnel

In a future implementation scenario, continuous monitoring during all sessions would be conducted by the researcher delivering the protocol, who would be trained in the recognition of physiological and behavioural indicators of distress in paediatric neurodevelopmental populations, with immediate access to a clinician affiliated with the host institution for safety escalation. All personnel responsible for session monitoring would be required to have received specific training in the recognition of physiological and behavioural indicators of distress, including autonomic changes, non-verbal dissent markers, and signs of sensory overload in behaviourally non-responsive children.Monitoring personnel would be authorised to interrupt or terminate sessions immediately if predefined safety thresholds were met or if any indication of discomfort or distress were observed, in accordance with the predefined stopping rules outlined in Table 1 and Section 4.5 as part of a pre-empirical safety governance framework.

### 4.3 Visual Mirror Exposure Protocol

The visual mirror exposure protocol described in this manuscript consists of brief, repeated, and strictly controlled sessions of passive visual stimulation. The protocol is designed to expose the child to a simple, coherent, and highly predictable visual environment, without requiring instruction following, task engagement, or voluntary behavioural responses. It is specifically adapted to behaviourally non-responsive paediatric populations.

#### Session Structure

In a prospective implementation, each session would comprise the following sequential phases, delivered in a fixed and predefined order:

##### 1. Baseline Phase

A resting baseline would be recorded prior to visual exposure. During this phase, no visual stimulus would be presented. Physiological signals would be recorded to establish the individual resting autonomic state.

##### 2. Stabilisation Phase

A static, high-contrast visual stimulus would be presented against a uniform background. This phase is intended to allow exposure to a stable visual signal without the need to process motion, change, or novelty.

##### 3. Continuity Phase

The visual stimulus would undergo slow, continuous, and regular motion, such as linear translation at a constant velocity. No acceleration, abrupt transitions, or unpredictable changes would be introduced.

##### 4. Controlled Novelty Phase

Limited and predictable modifications would be introduced to the stimulus, such as gradual changes in colour, size, or trajectory. These modifications would be predefined, infrequent, and temporally constrained, and would be intended to probe autonomic sensitivity while minimising the risk of sensory overload.

All phases would be delivered in a fixed sequence. No adaptive or contingent modifications would be applied within a session.

#### Stimulus Parameters

Across all sessions, visual stimuli would be characterised by:

- A very limited number of objects presented simultaneously;
- High contrast between stimulus and background;
- Slow and continuous motion when movement is present;
- Absence of faces, social content, symbolic material, or rapid visual events.

Stimulus parameters, including duration, luminance, contrast, colour range, and motion characteristics, would be predefined and would remain identical across participants, except where minor adjustments might be required to ensure tolerability.

#### Temporal Progression

In a future empirical context, the protocol would follow a predefined temporal progression across multiple weeks. Gradual increases in perceptual complexity would be introduced only after successful completion of earlier phases, as defined by the absence of predefined physiological or behavioural stopping criteria.

Early sessions would prioritise static and highly predictable visual stimuli. Subsequent sessions would introduce slow motion and, later, controlled and limited novelty in a structured and non-adaptive manner. This progression is intended to minimise perceptual load and prediction error while allowing repeated exposure under stable and controlled conditions.

The full session schedule, phase composition, and temporal progression would be specified a priori and would be summarised in Table 2 as an illustrative implementation example.

**Table 2.** Illustrative Visual Mirror Exposure Protocol: Session Structure, Stimulus Parameters, and Temporal Progression (Pre-Empirical Framework).

| Phase | Visual content | Motion characteristics | Duration (minutes) | Predictability level | Purpose |
| --- | --- | --- | --- | --- | --- |
| Baseline | No stimulus | None | 3 | Not applicable | Illustratively intended to establish an individual autonomic resting state. |
| Stabilisation | Single high-contrast static object | None | 4 | Maximal | Illustratively intended to promote visual stability and reduce sensory load. |
| Continuity | Same object | Slow linear motion at constant velocity | 4 | High | Illustratively intended to support temporal predictability and sustained engagement. |
| Controlled novelty | Same object | Predefined gradual change (colour, size, or trajectory) | 2 | Moderate, predefined | Illustratively intended to probe autonomic sensitivity without sensory overload. |

| Week range | Phases included | Novelty introduction |
| --- | --- | --- |
| Weeks 1 to 2 | Baseline, Stabilisation | None |
| Weeks 3 to 4 | Baseline, Stabilisation, Continuity | None |
| Weeks 5 to 6 | All phases | Controlled novelty only |

Note. The session structure, stimulus parameters, and temporal progression presented in this table are provided as illustrative, pre- empirical examples intended to inform the methodological design of future feasibility studies. They do not correspond to sessions delivered, participants exposed, or procedures implemented in an ongoing study.

Table 2 summarises the structure of the visual mirror exposure protocol as an illustrative, pre-empirical framework, including session phases, stimulus parameters, and temporal progression. All durations, stimulus characteristics, and phase sequences are specified a priori to illustrate protocol stability and safety considerations, rather than to describe procedures applied in an implemented study. The protocol is intentionally brief to minimise physiological burden and to ensure tolerability in a behaviourally non-responsive paediatric population. Temporal progression across weeks is fixed at the design level and is not intended to adapt to observed responses, except conceptually when predefined safety stopping rules would be triggered in a future implementation. This table provides the operational specification at the protocol design stage required for methodological transparency and ethical oversight.

#### Protocol Integrity

Within a prospective implementation, all sessions would be delivered according to the predefined protocol without modification based on observed physiological responses, except in cases where stopping rules would be triggered. Any deviation from the planned session structure, including early termination, would be documented and considered in feasibility assessment.

### 4.4 Outcomes and Measures

The present manuscript does not report an implemented feasibility study and does not aim to assess therapeutic efficacy or functional visual improvement.

All outcomes are conceptual, methodological, and exploratory, and are selected to define feasibility, tolerability, and measurement reliability criteria relevant to the visual mirror paradigm in a behaviourally non-responsive paediatric population.

Outcome measures are limited to non-behavioural physiological signals, defined as autonomic physiological measures that could be recorded without requiring voluntary motor, verbal, or task-based responses from the child. Autonomic physiological measures are not interpreted as specific markers of visual processing, but as non-behavioural indicators of physiological regulation, tolerability, and phase-dependent sensitivity to controlled visual input. These measures are chosen to allow conceptual examination of physiological engagement and tolerability during passive visual exposure.

#### Primary Outcomes

The primary outcomes defined here relate to physiological tolerability of the protocol. Tolerability is operationalised as the absence of sustained autonomic destabilisation indicative of distress during or following exposure. The following measures are identified as relevant indicators and would be recorded continuously throughout baseline, exposure, and post-exposure phases in a future empirical study:

- Heart rate, used to detect acute or sustained autonomic activation relative to individual baseline.
- Heart rate variability, used as an index of autonomic flexibility and parasympathetic regulation.

Predefined thresholds for physiological alert and stopping rules are specified at the design stage and detailed in Section 4.5.

#### Secondary Outcomes

Secondary outcomes are methodological and descriptive in nature. They include:

- The feasibility of acquiring continuous, interpretable physiological signals across all session phases.
- The presence or absence of non-verbal dissent markers, defined as behavioural or physiological signals that would be interpreted as refusal or discomfort, including abrupt increases in heart rate, sustained reduction in heart rate variability, sudden motor stiffening, agitation, or atypical vocalisations.
- The proportion of sessions that would be expected to be completed as planned versus interrupted or terminated early due to safety criteria.
- Intra-individual stability and variability of physiological signals across repeated sessions.

No outcome relies on behavioural performance, visual choice, task completion, or explicit communication from the child.

#### Measurement Windows

Physiological measures would be analysed within predefined temporal windows corresponding to each session phase. Baseline measures would serve as the individual reference state. Changes observed during stabilisation, continuity, and controlled novelty phases would be interpreted relative to this baseline, without inference of cognitive or perceptual content.

Periods affected by signal artefact, defined as loss or distortion of the physiological signal rendering data uninterpretable due to excessive movement or sensor displacement, would be flagged and excluded according to predefined criteria.

All outcomes and indicators are summarised in Table 3 as part of the proposed methodological framework.

**Table 3.** Illustrative outcome domains, measures, indicators, and interpretation framework (pre-empirical).

| Outcome domain | Measure | Phase(s) | Indicator | Interpretation purpose |
| --- | --- | --- | --- | --- |
| Physiological Tolerability | HR | All phases | Stability relative to baseline | Illustratively intended to inform the assessment of autonomic tolerability of exposure without inference of therapeutic effect. |
| Physiological Tolerability | HRV | All phases | Maintenance of autonomic flexibility | Illustratively intended to inform the detection of physiological stress or destabilisation as safety indicators only. |
| Feasibility of Measurement | Signal Acquisition Quality | All phases | Proportion of usable signal | Illustratively intended to inform the feasibility of physiological monitoring in a non-responsive population. |
| Non-Verbal Dissent | Behavioural and Physiological Markers | All phases | Presence of predefined indicators | Illustratively intended to define conditions under which predefined stopping rules would be triggered as an ethical safeguard, not as an outcome measure. |
| Session Completion | Session Status | Whole session | Completed versus interrupted | Illustratively intended to inform the assessment of procedural feasibility independent of clinical change. |
| Measurement Reliability | Intra-individual signal patterns | Across sessions | Signal consistency | Illustratively intended to inform the evaluation of methodological robustness without group-level inference. |

**Note:** HR: Heart Rate; HRV: Heart Rate Variability
The outcomes, measures, indicators, and interpretation purposes presented in this table are provided as illustrative, pre-empirical elements intended to inform the design and interpretation framework of future feasibility studies. They do not correspond to data collected, analyses performed, or outcomes evaluated in an implemented study.

Table 3 summarises the predefined outcomes, measures, and indicators as part of a pre-empirical methodological framework intended to inform the evaluation of feasibility, tolerability, and measurement reliability in future feasibility studies. All outcomes are specified a priori and are strictly non-behavioural in nature. The structure is designed to conceptually examine whether stable and interpretable physiological data could be collected during passive visual mirror exposure, while ensuring participant safety. These outcomes are not intended to support claims regarding visual perception, cognitive processing, or therapeutic efficacy, but to establish the methodological preconditions required for future mechanistic or interventional studies.

### 4.5 Safety and Stopping Rules

In a prospective implementation, participant safety would be ensured through predefined physiological and behavioural stopping rules, operationalised in Table 1 and applied as part of a pre-empirical safety governance framework. The visual mirror exposure protocol would be governed by these predefined safety thresholds and stopping rules, which are designed to prioritise physiological and behavioural indicators of potential discomfort or distress in a behaviourally non-responsive paediatric population.

#### Safety Monitoring Procedures

In a future empirical context, safety monitoring would be conducted continuously during all session phases by trained personnel, with immediate authority to interrupt the session if predefined stopping criteria were met. Monitoring would be performed in real time throughout baseline, exposure, and post-exposure periods.

The absence of overt behavioural response would not be interpreted as consent or tolerance. Instead, safety assessment would be based on objective physiological changes and observable behavioural signs that may indicate non-verbal dissent.

Figure 1 provides an illustrative, pre-empirical example of how real-time physiological safety monitoring could be implemented during a single visual mirror exposure session, including the definition of an individual baseline and the application of predefined safety thresholds. This figure is provided for conceptual clarification only and does not represent empirical data.

**Figure 1.**
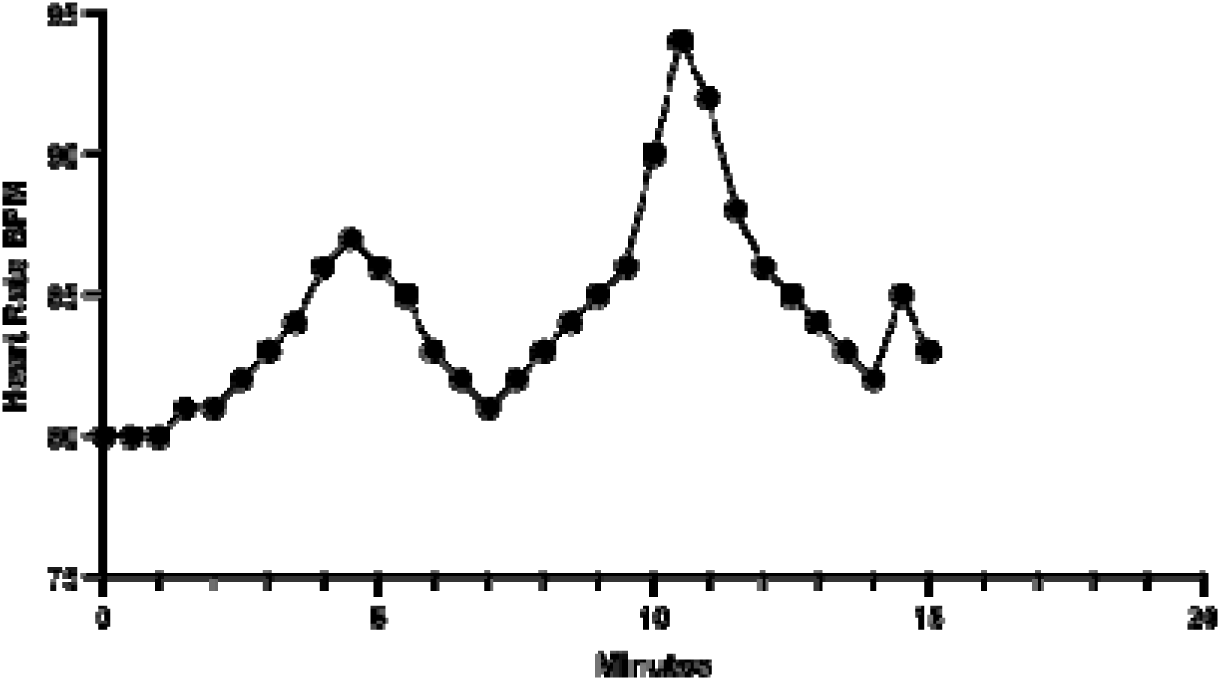
Illustrative, pre-empirical example of real-time physiological safety monitoring during a single visual mirror exposure session (mock-up).

This illustrative mock-up depicts how physiological safety monitoring could be implemented during a passive visual mirror exposure session. The solid grey horizontal line represents the individual resting heart rate baseline. The dashed red line indicates the predefined safety threshold corresponding to a +20% increase relative to baseline. The shaded area represents the predefined physiological safety zone. The solid line represents a simulated example of heart rate variation over time across session phases, including a controlled novelty phase. In this illustrative scenario, physiological modulation remains within predefined safety limits, and no stopping criteria would be triggered. This figure is provided for explanatory purposes only and does not represent empirical data.

#### Physiological Stopping Criteria

In a prospective implementation, sessions would be immediately terminated if any of the following physiological criteria were met:

- A sudden and sustained increase in heart rate exceeding approximately 20% above the individual resting baseline, persisting beyond transient fluctuations;
- A marked and persistent reduction in heart rate variability relative to baseline levels, indicative of reduced autonomic flexibility;
- Any pattern of autonomic destabilisation suggesting sustained physiological activation rather than transient orienting responses.

These thresholds are defined as operational precautionary criteria, rather than diagnostic markers. They are specified a priori to prevent prolonged exposure to potential stress in the absence of subjective reporting.

#### Behavioural Stopping Criteria

In parallel with physiological monitoring, sessions would be terminated immediately if any of the following behavioural indicators were observed:

- Sudden motor stiffening or freezing not present at baseline;
- Marked agitation, repetitive movements, or abrupt changes in posture;
- Atypical vocalisations or changes in breathing pattern suggestive of distress;
- Any behavioural change that would be judged by the supervising clinician or researcher to reflect discomfort or refusal.

Behavioural indicators would be interpreted conservatively. When uncertainty exists, the decision rule would systematically favour immediate session termination.

#### Stopping Procedures and Documentation

If a stopping criterion were met, visual stimulation would be ceased immediately, and the child would be returned to a neutral resting state. No attempt would be made to resume the session on the same day.

All instances of early termination, including the triggering criterion and session phase, would be documented and would be incorporated into feasibility and tolerability considerations. Early termination would not be considered a protocol deviation but a predefined indicator relevant to feasibility assessment.

#### Ethical positioning

These stopping rules operationalise a precautionary ethical governance approach in which participant protection supersedes data completeness. These stopping procedures are not intended to infer subjective emotional states, but to provide clear, auditable documentation of discontinuation decisions based on observable biological and behavioural signals.

The presence, frequency, and context of stopping events are defined here as conceptual feasibility indicators, intended to inform the ethical viability of future empirical investigations using this paradigm.

### 4.6 Procedure timeline

In a future empirical context, the procedure would follow a predefined and fixed timeline designed to evaluate feasibility, tolerability, and measurement reliability of the visual mirror paradigm across repeated exposures. The timeline is structured to minimise sensory load, ensure participant safety, and allow conceptual examination of intra-individual physiological stability over time.

Each participant would follow the same temporal progression, independent of physiological responses, except when predefined stopping rules were triggered. No adaptive modification of stimulus parameters would be performed based on interim observations. No interim analyses or protocol adaptations would be planned.

The protocol would be organised into three successive phases spanning six weeks. Progression from one phase to the next would follow a predefined fixed schedule and would not be adapted based on observed responses, except in cases where predefined stopping rules would result in session termination. Successful completion would be defined retrospectively for feasibility considerations and would not alter the planned timeline.

Early sessions would prioritise maximal predictability and perceptual stability. Subsequent sessions would introduce controlled and limited novelty in order to probe autonomic sensitivity while maintaining a low cognitive and sensory burden.

All sessions would be scheduled with sufficient temporal spacing to avoid cumulative fatigue or sensitisation effects. Session frequency and duration would be identical across participants.

The full procedure timeline, including session content and duration, is specified a priori at the design stage and is summarised in Table 4 as an illustrative implementation example.

**Table 4.**
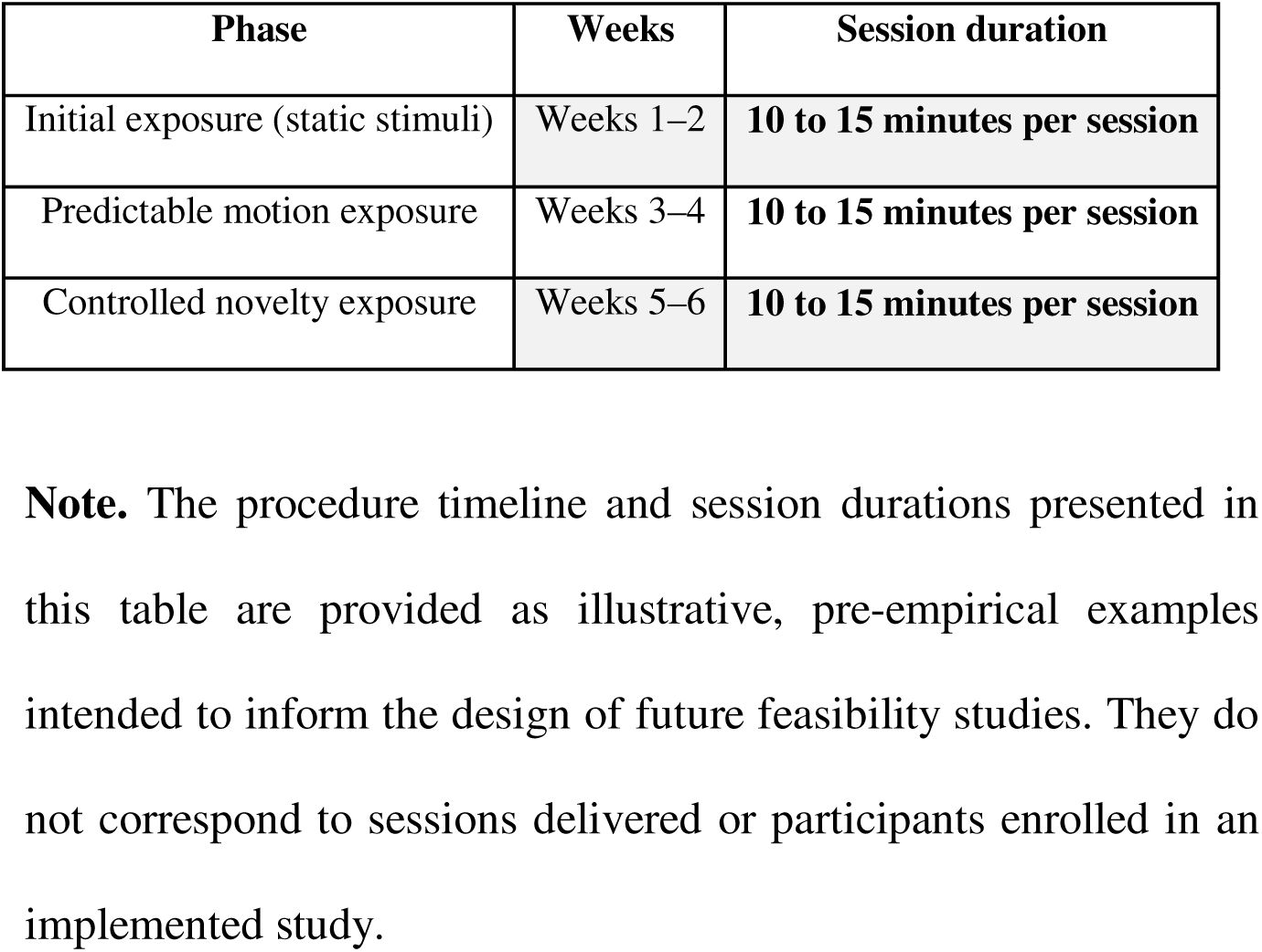
Illustrative procedure timeline and visual mirror exposure progression (pre-empirical framework).

Table 4 summarises the predefined temporal structure of the visual mirror exposure protocol as an illustrative, pre-empirical framework. Each phase is designed to incrementally modulate perceptual complexity while preserving high stimulus predictability.

Session durations are intentionally brief to reduce physiological and sensory burden. The progression from static to dynamic stimuli, followed by controlled novelty, reflects the theoretical aim of gradually engaging visual predictive mechanisms without inducing overload.

This timeline does not represent a therapeutic dose escalation and is not intended to induce functional change. Rather, it is intended to illustrate how protocol feasibility, tolerability, and physiological signal stability could be examined within a predefined, feasibility-focused design framework, prior to any empirical implementation.

## 5. Data Analysis Plan

This section outlines a predefined, pre-empirical descriptive analysis framework intended to inform the evaluation of feasibility, tolerability, and measurement reliability of the visual mirror paradigm in future feasibility studies. The analyses focus exclusively on non-behavioural physiological signals, predefined temporal windows corresponding to protocol phases, and explicit decision rules specified a priori. No hypothesis testing, efficacy evaluation, or inferential analysis is proposed at this stage.

The complete data analysis plan, including signals, temporal windows, indicators, and decision thresholds, is specified at the protocol design stage and is summarised in Table 5 as an illustrative analytical framework.

**Table 5.** Illustrative data analysis framework: signals, temporal windows, indicators, and decision rules (pre-empirical).

| **Analysis domain** | **Physiological signal** | **Temporal window** | **Descriptive metric** | **Indicator** | **Decision rule** |
| --- | --- | --- | --- | --- | --- |
| **Baseline Physiological State** | HR | Pre-exposure | Mean HR | Stable autonomic baseline | Baseline would be considered valid if the signal were continuous and artefact-free at the individual level. |
| **Baseline Physiological State** | HRV | Pre-exposure | Time-domain indices | Interpretable baseline | Baseline indices would be retained if they were computable at the individual level. |
| **Exposure Tolerability** | HR | Stabilisation | Mean and peak HR | Absence of sustained activation | Session continuation would be supported if HR were to remain within predefined thresholds at the individual level |
| **Exposure Tolerability** | HRV | Stabilisation | Qualitative pattern | Autonomic stability | Session interruption would be considered if a sustained reduction indicative of distress were to be observed. |
| **Sensitivity to Controlled Novelty** | HR | Controlled novelty phase | Peak and pattern relative to baseline | Autonomic reactivity | Data inclusion would be considered if signal quality were to meet the criteria specified in Table 1; session termination would be considered if predefined non-verbal dissent markers (Section 4.5) were to be detected. |
| **Non-Verbal Dissent** | Combined Signals | All phases | Presence of markers | Ethical safety | Immediate termination would be considered upon detection. |
| **Measurement Reliability** | HR and HRV | Across sessions | Intra-individual consistency | Signal stability | Descriptive assessment would be conducted at the intra-individual level across repeated sessions. |
| **Feasibility** | Procedural | Entire protocol | Completion rate | Implementability | Overall feasibility would be evaluated descriptively at the |
| **Assessment** | Indicators |  |  |  | protocol design level, using an illustrative feasibility sample size (e.g. n = 10), without group-level inference. |

Note: HR: Heart Rate; HRV: Heart Rate Variability
The analysis domains, signals, temporal windows, indicators, and decision rules presented in this table are provided as illustrative,
pre-empirical elements intended to inform the analytical framework of future feasibility studies. They do not correspond to analyses
conducted, decisions applied, or data generated in an implemented study.

Table 5 summarises the predefined data analysis plan as part of a pre-empirical methodological framework, linking each analytical domain to specific physiological signals, temporal windows, descriptive metrics, and explicit decision rules. In line with the objectives of a feasibility-focused study, the table specifies how physiological data could be examined descriptively at the individual level, without hypothesis testing or group-level inference. The decision rules outlined in the table define a priori the conditions under which protocol feasibility, physiological tolerability, and measurement reliability would be considered achieved, partially achieved, or not achieved. By specifying these analytical procedures at the protocol design stage, the table ensures transparency, prevents analytical flexibility, and establishes a clear interpretive structure for interpreting outcomes strictly in terms of feasibility rather than therapeutic efficacy.

### 5.1 General Analytical Structure

This manuscript does not report an implemented feasibility study. It is not designed to test therapeutic efficacy, causal effects, or group-level hypotheses.

All analyses described here are pre-specified at the protocol design level and are intended to inform future feasibility studies. They are descriptive, exploratory, and strictly limited to the evaluation of feasibility, tolerability, and measurement reliability of the visual mirror paradigm.

Analyses would focus on intra-individual trajectories rather than between-participant comparisons. Each child would serve as their own reference, and no inferential statistics aimed at generalisation would be planned.

No hypothesis testing, effect size estimation, or correction for multiple comparisons is proposed.

### 5.2 Data Preprocessing and Signal Quality Control

Physiological signals would be visually and algorithmically inspected prior to analysis to identify artefacts related to movement, sensor displacement, or technical failure.

Segments affected by signal artefact would be flagged and excluded according to predefined criteria. Only periods with interpretable physiological data would be retained for analysis.

Baseline values would be computed individually for each participant using resting-state segments recorded prior to visual exposure in a future implementation.

### 5.3 Descriptive Analysis of Physiological Signals

Heart rate and heart rate variability would be summarised descriptively within predefined temporal windows corresponding to protocol phases.

Analyses would examine:

- baseline resting periods,
- stabilisation phases,
- continuity phases,
- controlled novelty phases.

For each participant, temporal evolution of physiological signals would be visualised across sessions using within-subject plots and time-series representations.

No aggregation across participants would be performed.

### 5.4 Assessment of Feasibility and Tolerability

Feasibility outcomes would be evaluated using procedural indicators, including:

- number of sessions initiated,
- number of sessions completed,
- frequency and timing of early session termination,
- occurrence of protocol deviations.

Tolerability would be assessed through the absence or presence of sustained physiological activation exceeding predefined thresholds, and through documentation of non verbal dissent markers triggering session interruption.

These outcomes would be summarised descriptively at the individual level.

### 5.5 Assessment of Measurement Reliability

Measurement reliability would be examined by evaluating:

- stability of physiological signals across repeated sessions within individuals,
- consistency of signal patterns across comparable protocol phases,
- proportion of sessions yielding usable physiological data.

Reliability assessment does not assume stationarity or normality of signals and would remain limited to descriptive pattern identification.

### 5.6 Interpretation Structure Specific to Feasibility Assessment

Interpretation would rely on within-session and within-individual contrasts across predefined protocol phases, rather than on absolute autonomic values.

Differential autonomic patterns would be used to characterise the internal structure and interpretability of physiological signals. They would not be used to infer magnitude of response, functional visual processing, or therapeutic effect.

Outcomes would be interpreted as indicating:

- feasibility achieved,
- feasibility partially achieved,
- feasibility not achieved,

Based on the combination of protocol completion rates, physiological tolerability, and signal interpretability at the individual level.

The predefined target sample size of 10 participants is specified as an illustrative feasibility objective, rather than a rigid requirement. No post hoc modification of decision thresholds would be permitted.

Importantly, the protocol would be considered feasible even if physiological stabilisation reflects habituation rather than engagement, provided predefined tolerability and signal interpretability criteria are met.

### 5.7 Transparency and Reproducibility

All analysis procedures, decision rules, and feasibility criteria are defined a priori at the protocol design stage, prior to any data collection, to minimise analytical flexibility.

Any deviation from the planned analytical procedures would be documented and reported alongside feasibility considerations in a future implementation.

## 6. Ethics, Oversight, and Regulatory Considerations

This manuscript describes a clinical feasibility study protocol and does not involve the collection, analysis, or reporting of empirical data from human participants.

No study procedures have been initiated, and no participants have been recruited.

The proposed visual mirror paradigm is intended for future empirical implementation in a highly vulnerable paediatric population. Consequently, ethical considerations are integrated at the protocol design level, prior to any data collection. Formal ethical approval will be sought from the appropriate Human Research Ethics Committee at the host institution before the commencement of any study involving human participants.

The protocol is designed in accordance with the precautionary principle. The visual mirror exposure is strictly passive, non-invasive, and does not require instruction following, task engagement, or voluntary behavioural responses. The absence of overt behavioural response is not interpreted as consent. Participant protection relies on continuous monitoring of predefined physiological and behavioural indicators of potential non-verbal dissent, including autonomic destabilisation and observable signs of distress. Any indication of predefined non-verbal dissent would result in immediate termination of the session, in accordance with the stopping rules specified in Section 4.5.

No therapeutic intent is associated with this protocol. The study does not aim to induce functional change, does not replace or withhold standard care, and does not involve deception, randomisation, or experimental manipulation beyond controlled visual exposure. The sole purpose of the proposed protocol is to determine whether the paradigm can be implemented safely, tolerably, and with sufficient measurement reliability to justify consideration of future mechanistic or efficacy studies.

Data management, storage, and analysis procedures for any future implementation will comply with institutional, national, and international regulations governing research involving children and vulnerable populations. In a future empirical implementation, the protocol could be discontinued if predefined feasibility, safety, or tolerability criteria are not met. Such an outcome would be considered ethically appropriate and informative for future research planning.

## 7. Discussion

The present manuscript describes a clinical feasibility study protocol designed to evaluate a visual mirror paradigm in children with cerebral visual impairment (CVI) presenting an atelic profile. As no empirical data have been collected, this discussion does not interpret outcomes or effects, but instead examines the methodological validity, scope, and limits of the proposed protocol. The primary methodological value of this protocol lies in its explicit adaptation to a population for whom conventional visual assessment or rehabilitation approaches are neither ethically nor practically applicable. By relying exclusively on passive visual exposure and non-behavioural autonomic physiological measures, the protocol avoids any requirement for voluntary responses, task engagement, or instruction following. This design directly addresses a major methodological gap in the study of children with severe CVI and atelic profiles, while remaining compatible with stringent ethical constraints.

A central strength of the proposed approach is its exclusive focus on feasibility, tolerability, and measurement reliability, rather than on clinical efficacy or functional visual processing. The use of predefined session phases and within-session, within-individual contrasts allows evaluation of whether physiological signals exhibit structured and interpretable organisation in response to controlled visual input, without reliance on absolute autonomic values. This structure is specifically intended to determine whether the proposed paradigm is sufficiently stable, interpretable, and ethically manageable to justify future empirical implementation.

The protocol also incorporates conservative safety monitoring and stopping rules designed to protect participants who may be unable to express discomfort through overt behavioural or verbal means. By clearly distinguishing between analytic inclusion criteria and ethical termination criteria, the design prioritises participant protection while preserving methodological transparency. These safeguards are essential when working with highly vulnerable paediatric populations and constitute a core component of the feasibility assessment.

Several limitations inherent to the proposed design must be acknowledged. The absence of behavioural outcomes, group-level comparisons, or functional performance measures necessarily restricts the interpretive scope of the data that may be generated by future implementation. In addition, autonomic physiological signals are not interpreted as specific markers of visual processing, but rather as indirect indicators of physiological regulation, tolerability, and signal stability. These constraints are intentional and reflect the ethical and practical boundaries associated with research involving children with severe cerebral visual impairment and atelic profiles, rather than methodological shortcomings.

The predefined feasibility, tolerability, and measurement reliability criteria are intended to guide both the ethical conduct and interpretation of the study. Should these criteria not be met, early discontinuation of the protocol would constitute an ethically appropriate and informative outcome. Conversely, if the criteria are satisfied, the findings may inform future methodological or clinical research considerations, without implying any obligation or commitment to subsequent studies.

This discussion therefore addresses the validity and limits of the proposed methodological approach itself, rather than interpreting outcomes that have not yet been observed. The manuscript should be read as a methodological protocol evaluated prior to data collection, establishing clear ethical, procedural, and analytical boundaries for any future empirical investigation.

## Declarations

### Ethics Statement

This manuscript presents a conceptual and methodological protocol developed within an independent research programme. No data involving human participants were collected, generated, or analysed as part of the present work.

The proposed protocol is intended to inform future empirical research involving vulnerable paediatric populations. Formal ethical approval will be sought from the appropriate Human Research Ethics Committee (HREC) at the host institution prior to the commencement of any study involving human participants.

The protocol has been designed in accordance with the precautionary principle. Given the limited capacity for verbal assent in the target population, the absence of a behavioural response is not interpreted as consent. The protocol explicitly integrates physiological and behavioural indicators as markers of potential non-verbal dissent, and any such indicators would lead to immediate termination of the session in future implementations.

Generative AI tools were used during the preparation of the manuscript to facilitate linguistic revision from French to academic English. This tool was not used to generate scientific content, interpret data, or draw conclusions. All content has been reviewed, verified, and approved by the author.

## Data and Materials Availability

No empirical data were collected for this study. The protocol materials and methodological documentation have been archived in a private repository and can be shared with reviewers or editors upon request.

## Data Availability Statement

Not applicable. No data were generated or analysed in the current study.

